# HIV self-testing positivity rate and linkage to confirmatory testing and care: a telephone survey in Côte d’Ivoire, Mali, and Senegal

**DOI:** 10.1101/2023.06.10.23291206

**Authors:** Arsène Kouassi Kra, Arlette Simo Fotso, Nicolas Rouveau, Mathieu Maheu-Giroux, Marie-Claude Boily, Romain Silhol, Marc d’Elbée, Anthony Vautier, Joseph Larmarange, the ATLAS team

## Abstract

HIV self-testing (HIVST) empowers individuals to decide when and where to test and with whom to share their results. From 2019 to 2022, the ATLAS program distributed ∼ 400 000 HIVST kits in Côte d’Ivoire, Mali, and Senegal. It prioritised key populations, including female sex workers and men who have sex with men, and encouraged secondary distribution of HIVST to their partners, peers and clients.

To preserve the confidential nature of HIVST, use of kits and their results were not systematically tracked. Instead, an anonymous phone survey was carried out in two phases during 2021 to estimate HIVST positivity rates (phase 1) and linkage to confirmatory testing (phase 2). Initially, participants were recruited via leaflets from March to June and completed a sociobehavioural questionnaire. In the second phase (September-October), participants who had reported two lines or who reported a reactive result were recontacted to complete another questionnaire. Of the 2 615 initial participants, 89.7% reported a consistent response between the number of lines on the HIVST and their interpretation of the result (i.e., ‘non-reactive’ for 1 line, ‘reactive’ for 2 lines).

Overall positivity rate based on self-interpreted HIVST results was 2.5% considering complete responses, and could have ranged from 2.4% to 9.1% depending on the interpretation of incomplete responses. Using the reported number of lines, this rate was estimated at 4.5% (ranging from 4.4% to 7.2%). Positivity rates were significantly lower only among respondents with higher education. No significant difference was observed by age, key population profile, country or history of HIV testing.

The second phase saw 78 out of 126 eligible participants complete the questionnaire. Of the 27 who reported a consistent reactive response in the first phase, 15 (56%, 95%CI: 36 to 74%) underwent confirmatory HIV testing, with 12 (80%) confirmed as HIV-positive, all of whom began antiretroviral treatment.

The confirmation rate of HIVST results was fast, with 53% doing so within a week and 91% within three months of self-testing. Two-thirds (65%) went to a general public facility, and one-third to a facility dedicated to key populations.

The ATLAS HIVST distribution strategy reached people living with HIV in West Africa. Linkage to confirmatory testing following a reactive HIVST remained relatively low in these first years of HIVST implementation. However, if confirmed HIV-positive, almost all initiated treatment. HIVST constitutes a relevant complementary tool to existing screening services.

## Introduction

Early testing followed by successful linkage to antiretroviral treatment for those diagnosed with HIV can drastically reduce the risk of onward HIV transmission and mortality [1–6]. In 2021, according to the United Nations Program for HIV/AIDS (UNAIDS), 81% of the adult population living with HIV in West and Central Africa knew their status. Only 77% of them were on antiretroviral treatment[7], below the 95-95-95 UNAIDS targets for 2025 [8]. The 95-95-95 targets aim for 95% of people living with HIV to know their status, 95% of those diagnosed to receive treatment, and 95% of those on treatment to achieve viral suppression. Improving diagnosis coverage, especially among vulnerable key populations at high risk of HIV acquisition and transmission, is the necessary first step to achieve this goal.

HIV self-testing (HIVST) is the process by which a person who wants to know their HIV status collects their own sample (oral fluid or blood), performs the test, and then interprets the results themself, often in a private setting [9]. It is an innovative tool that empowers individuals and guarantees the confidentiality of the test result [10]. Individuals may decide when and where to test and with whom they want to share their result. It is a tool that is widely accepted by various populations, including key populations [11–18]. It has been shown to be effective in screening populations vulnerable to HIV acquisition and transmission that are often hardly reached through conventional approaches [19–21]. The World Health Organization (WHO) has recommended HIVST as a complementary testing approach since 2016 [22].

The HIV Self-Testing in Africa (STAR) project carried in Eastern and Southern Africa and funded by Unitaid aimed to boost the global market for HIVST (https://www.psi.org/fr/project/star/). The project unfolded in three phases: Phase 1 ran from September 2015 to August 2017, Phase 2 spanned from August 2017 to July 2020, and Phase 3 took place between January 2020 and July 2021. Following the experience gained in Eastern and Southern Africa under the STAR project [11, 23–28], the Unitaid funding agency sought to stimulate HIVST in West Africa, where HIV epidemics are distinguished by their more concentrated and less generalised nature compared to those in Eastern and Southern Africa. In this region, the general population prevalences are relatively low to very low, and key populations (for example, female sex workers and men who have sex with men) are particularly affected and bear a disproportionate share of the HIV burden [29]. The ATLAS programme (*AutoTest de dépistage du VIH : Libre d’Accéder à la connaissance de son Statut*) aimed to promote, implement, and expand HIVST in Côte d’Ivoire, Mali, and Senegal [30] where the national HIV prevalence in 2021 was 1.9% (1.7%-2.2%), 0.8% (0.6%-1%), and 0.3% (0.3%-0.4%) respectively [31].

To preserve the anonymity and confidentiality of HIVST and not impede their use, ATLAS decided, in line with WHO recommendations, not to track the use and outcomes of distributed HIVST kits systematically. Such tracking can be logistically challenging and costly and could limit the distribution, redistribution and use of HIVST [32]. Without systematic tracking, it is challenging to obtain information on the users of the HIVST, their results and on linkage to confirmatory testing and treatment, which are crucial indicators to assess program effectiveness and impact. For instance, the positivity rate can reflect the yield of new individuals diagnosed with HIV and if the testing modality is indeed reaching those in need. Diagnosed individuals must seek confirmatory testing and be linked to care to maximise health benefits and decrease onward transmission.

We conducted an innovative survey by setting up an anonymous and free telephone platform in Côte d’Ivoire, Mali and Senegal while preserving anonymity and encouraging voluntary participation. In the second phase (September-October), participants who had reported two lines or a self-interpreted HIVST result as reactive were recontacted to complete another questionnaire. Here we present the HIV test positivity rates from the phase 1 questionnaire and the links with confirmatory tests and care.

## Materials and Methods

### ATLAS program description

ATLAS HIVST distribution was integrated into existing testing policies, programmes and activities in each country; 397 367 HIVST kits were freely distributed between July 2019 and February 2022 as part of the three countries’ national AIDS strategies. At the time of ATLAS’s implementation in 2019, only small-scale HIVST pilot programs had been previously conducted in Senegal and Côte d’Ivoire, whereas Mali had no previous experience with HIVST. In Senegal, for instance, the first pilot survey took place between April 2017 and June 2018 [33].

The design of the different delivery channels and the priority populations were developed with country stakeholders including national AIDS programs/councils, international institutions including the WHO, international and national non-governmental organisations involved in local HIV programs, and civil society and community leaders. ATLAS HIVST distribution was organised through eight different operational delivery channels (Figure 1), i.e. five facility-based approaches (delivery of HIVST kits through public or community-based health facilities) and three community-based approaches involving outreach activities engaging female sex workers (FSW), men who have sex with men (MSM), and people who use drugs (PWUD) [30]. Peer educators conducted these outreach activities through group activities (e.g. talks, discussion groups, night visits, social events, or parties) and face-to-face activities (e.g. home visits). Outreach activities represented the majority (∼85%) of ATLAS’s distribution volume.

**Figure 1.**
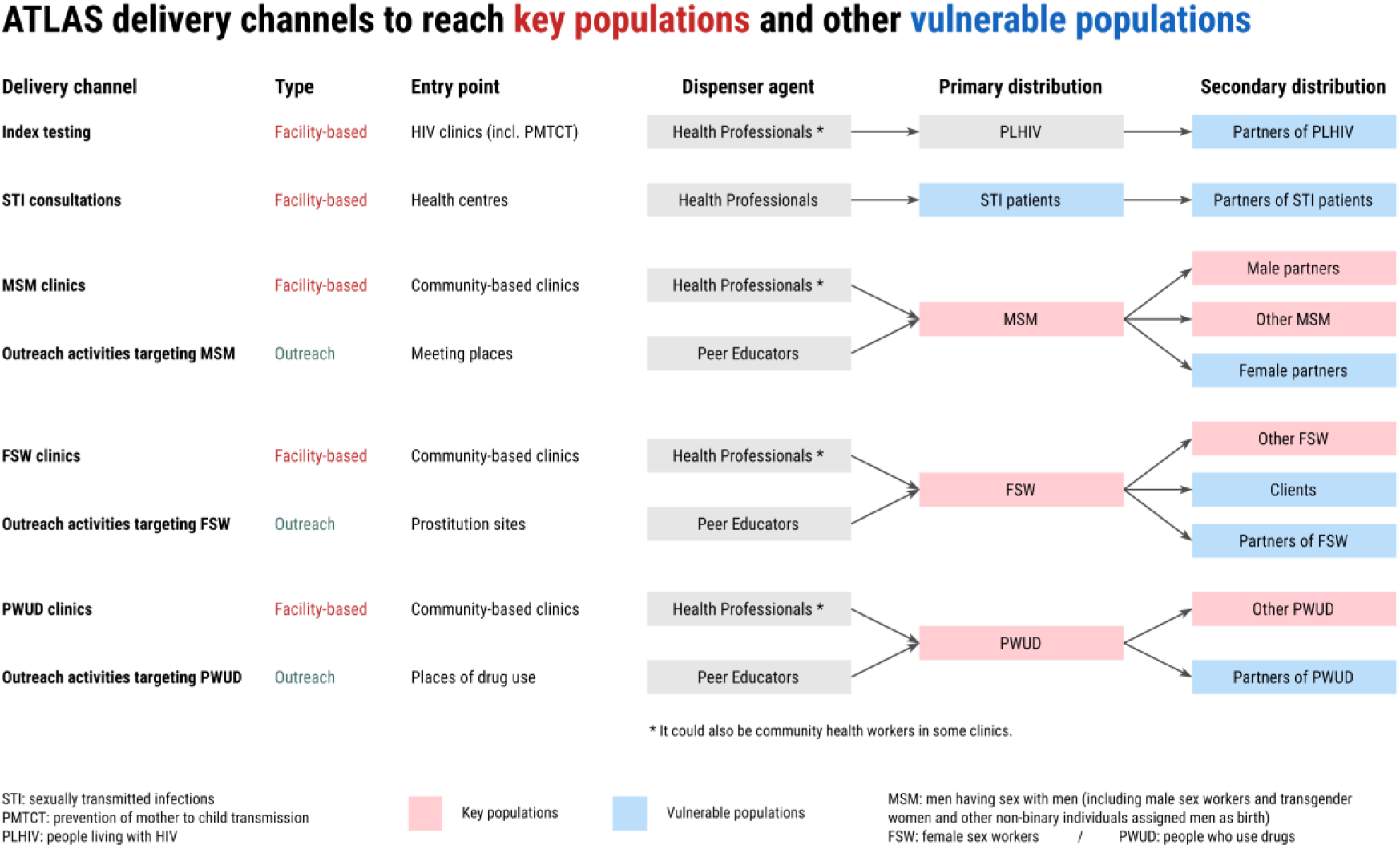
ATLAS delivery channels (adapted from [30]). FSW=female sex workers, MSM=men who have sex with men, PLHIV=people living with HIV PMTCT=prevention of mother-to-child transmission, PWUD=people who use drugs, STI=sexually transmitted infection.

ATLAS activities relied on both primary and secondary distribution. HIVST kits were distributed by peer educators and healthcare professionals to primary contacts for their personal use (primary distribution). With secondary distribution, primary contacts were provided HIVST kits and invited to redistribute them to their peers, partners, and clients. These secondary contacts were often members of key populations that can be more difficult to engage in HIV prevention, along with other peripheral vulnerable populations. This chain-referral distribution of HIVST implies that end-users were not limited to primary contacts.

Only oral self-testing (OraQuick HIV Self-Test®) has been distributed through ATLAS. OraSure Technologies, the manufacturer of the OraQuick test, accompanies each HIVST kit with a user manual for result interpretation. OraQuick HIVST results should be interpreted as follow: “reactive” (“positive”) if two lines (C & T) are visible (even barely), “non-reactive” (“negative”) if only the C (control) line is visible, and “invalid” if no line is visible or if only the T (test) line is visible. To be noted, the French version of the HIVST instructions distributed by ATLAS (Figure 2, Figure S1) used the wording “reactive” / “non-reactive” instead of “positive” / “negative” to qualify the HIVST result, following WHO vocabulary in their HIVST guidelines [20] as an HIVST is triage test and does not provide a definitive HIV-positive diagnosis. The questionnaire of the survey also used “reactive” / “non-reactive” wording (https://doi.org/10.5281/zenodo.11061878).

**Figure 2.**
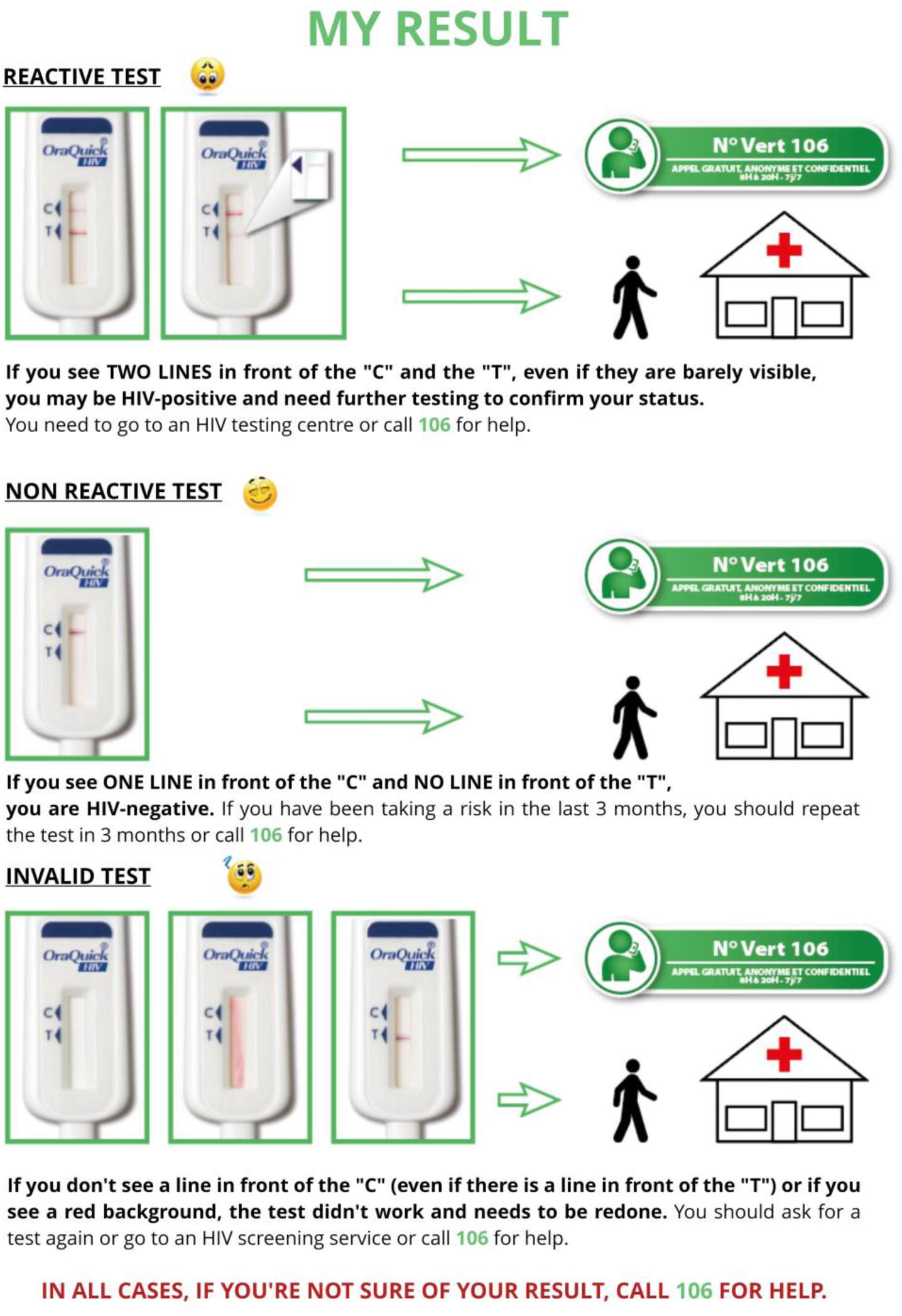
English translation of the guidelines for interpreting HIVST result, following manufacturer instructions for use (OraQuick HIV Self-Test®), as included in the ATLAS brochure distributed with HIVST (Ivorian version). See https://doi.org/10.5281/zenodo.11086135 for the original French version.

In addition to the manufacturer’s instructions, locally adapted brochures and explanatory videos in French and local languages have been developed to help users perform the test, interpret the result and know what actions should be taken following a non-reactive, a reactive or indeterminate result (for example : https://youtu.be/laCCjVEKZto in English or https://youtu.be/1xzitLD309U in French). They also encouraged people with a reactive HIVST to seek confirmatory HIV testing and care. Individuals with a non-reactive test were invited to retest after 3 months if still exposed to HIV. Existing toll-free hotlines in each counntry were strengthened and trained on HIVST, to offer information about HIV, prevention, testing, use and interpretation of HIVST and counseling.

### Study design and data collection

The ATLAS program embedded multiple research activities, from qualitative studies to economic analyses, which have been described in detail elsewhere [17, 30, 34–37].

The program included a voluntary anonymous phone survey. Between mid-March and mid-June 2021, dedicated survey flyers were distributed with the HIVST kits inviting self-test users in each country to call a toll-free number to complete a questionnaire (phase 1). All calls from the three countries, over the same period, were rerouted to a telephone platform located in Abidjan and operated by Ipsos Côte d’Ivoire, which was selected following an international call for tenders.

A pilot survey was initially conducted without offering financial compensation to the participants [38]. Following its results, we decided to introduce a reward as a token of appreciation for the time participants dedicated to the survey. Consequently, completion of the questionnaire was rewarded with 2 000 XOF (approximately 3.40 USD) of phone communication credit. This reward was mentioned on the survey flyers. In order to participate in the survey, participants had to be of legal age to use an HIVST on their own without parental permission (16 years in Côte d’Ivoire, 18 years in Mali, and 15 years in Senegal) and had to have used an HIVST provided to them through the ATLAS project.

As the survey was anonymous, there was a risk that some HIVST users may participate more than once or that individuals who have never used HIVST tried to participate to receive the financial incentive. To limit these risks, several measures were taken: (i) the leaflet distributed with the HIVST kits had a unique 9-digit number generated by the research team that was requested prior to participation in the survey, (ii) the same unique number could not be used twice, (iii) the financial incentive was only paid out once the questionnaire was fully completed (however individuals could refuse to answer any particular question), (iv) the same telephone number could not be used twice to receive the telephone credit. These unique 9-digit numbers were generated non-sequentially and were grouped by country, delivery channel and implementing partner. Thus, any unique number could indirectly identify the delivery channel where the HIVST kit was initially dispensed.

The time when participants received their HIVST kit was not collected. However, as a survey leaflet was mandatory to participate, we could estimate that all participants received their HIVST kit during the survey period (i.e. between mid-March and mid-June 2021).

The phase 1 questionnaire, which lasted 20 to 30 minutes, collected information on sociodemographic characteristics of HIVST users (including age, sex, marital status, education level), testing history (having ever tested for HIV before using HIVST and date of last HIV test), sexual and preventive behaviours, HIVST use and difficulties encountered [39]. Specifically, each participant was asked about the number of lines that appeared when reading the HIVST result and their self-interpretation of it (reactive or non-reactive).

In total, 2 615 participants were recruited for phase 1[39]. Those who reported two lines or a reactive result (n=126) were asked for their consent to be called back a few months later to participate in a complementary survey and, if consented, to provide a phone contact (n=120). As some individuals may delay their decision to undergo a confirmatory test by several weeks/months after using an HIV self-test, we chose a minimum of 3-month gap between our two surveys to potentially get an estimate of the maximum number of participants who eventually underwent confirmatory testing.

From September 27^th^ to October 22^nd^, 2021, 96 were successfully recontacted and invited to complete a 5-minute questionnaire (phase 2). Among those, 89 accepted to participate in phase 2 and 78 fully completed phase 2 questionnaire. Phase 2 questionnaire asked the participants if they had undergone a confirmatory test following their HIVST, reasons for not linking to confirmatory testing (if not linked), time between HIVST and confirmatory testing (if linked), type of facility for confirmatory testing, confirmation test result, linkage to antiretroviral treatment (if confirmed HIV-positive).

The interviews were conducted in either French, English, Bambara, or Wolof. On-the-fly translation into other local languages was also available. Compensation of XOF 2 000 (≈3.40 USD) in the form of telephone credit was given to participants who completed the phase 2 questionnaire. Unlike in phase 1, it was not a financial incentive as participants were informed about it only at the end of the interview. Interviews were not audio-recorded. Questionnaires’ responses were captured on a computer and stored in a database managed by PAC-CI, an Ivorian research institute with expertise in clinical research.

At the end of the survey, collected telephone numbers (for appointments and rewards) were deleted from the database. All procedures have been described in a publicly available data management plan (https://dmp.opidor.fr/plans/3354/export.pdf). The complete project protocol, including the data management plan (required by the ethics committees), was written in French. Both phase 1 and phase 2 questionnaires have been made available online and a link is now provided (https://doi.org/10.5281/zenodo.10210464).

### Data analysis

Following a previously published analysis [39], and due to the small numbers of participants in certain distribution channels, distribution channels (Figure 1) were grouped into three categories: FSW-based channels (outreach or facility-based), MSM-based channels (outreach or facility-based) and other channels (PWUD-based channels, index testing, STI consultations). As the profile of participants should differ substantially by sex and distribution channel (women from the FSW-based channel are more likely FSW while those from the MSM-based channel are more likely female partners of MSM; men from the MSM-based channel are more likely MSM while those from the FSW-based channel are more likely partners or clients of FSW, see Figure 1), we decided to combine distribution channel and sex into a single combined variable named key population profile.

Based on phase 1 participants’ self-reports, we distinguished between those who provided a consistent response, i.e. the reported number of visible lines was consistent with the reported self-interpretation (2 visible lines reported as reactive, one line reported as non-reactive, or no/one line and interpreted as invalid), those who provided an inconsistent response, i.e. the number of visible lines was inconsistent with the self-interpretation of the result, or those who returned only a partial response (refusal to answer or answered “I don’t know” to one or both questions).

Due to the inconsistency of responses, we considered self-reported results and reported number of HIVST lines separately to estimate HIVST positivity rates. For each source, we calculated positivity rates for complete responses (excluding ‘don’t know’ and refusals (DK-R) from the numerator and denominator). We also calculated the potential range of positivity rates by including incomplete responses (highest possible rate, DK-R responses are considered reactive, and lowest possible rate, DK-R responses are considered non-reactive).

We conducted two separate multivariable logistic regressions, based respectively on self-interpreted results and the reported number of lines, to analyse differences in positivity rates according to key population profile, country, age group, marital status, educational level, and first-time tester. Global p-values for each variable were computed using likelihood-ratio tests (using the *Anova()* function from ‘car’ R package). To account for multiple comparisons, q-values were computed with the Bonferroni correction (using the R *p.adjust()* function). We deemed it important to stratify the positivity rates by country, key population profile, and age group (15-24, 25-34, and 35+).

We described the selection of eligible participants for phase 2 questionnaires and corresponding participation rates. To assess any participation bias, characteristics of phase 2 participants (country, sex and distribution channel, age group, marital status, educational level, and first-time testers, i.e. whether they ever tested for HIV before using HIVST) were compared with individuals eligible for phase 2 but who did not participate and with phase 1 participants not eligible for phase 2. Simple comparisons were conducted using chi-square tests, and multiple comparison was performed using a multivariable multinomial logistic regression model, followed by the calculation of likelihood ratio tests.

Among phase 2 eligible participants who completed their questionnaire, linkage to confirmatory testing, the proportion being confirmed HIV positive, and the proportion who initiated treatment were described, stratified by the reported number of lines and self-interpreted HIVST result in phase 1 questionnaire.

We also described (i) for those who did not link to confirmatory testing, the main reported reason; and (ii) for those who did link to confirmatory testing, the type of facility attended for confirmation and the time between HIVST and confirmatory testing.

A dedicated anonymised dataset and the corresponding R script are available on Zenodo (https://doi.org/10.5281/zenodo.11086135) to allow replication of the analysis. All analyses have been performed using R version 4.3.1 [40]. All the descriptive tables were generated using the *tbl_summary()* function from the ‘gtsummary’ package [41]. Confidence intervals (95% confidence interval, 95%CI) were computed using Wilson’s method with Yate’s continuity correction (*prop.test()* function in the ‘stats’ package). Multinomial models were computed with *multinom()* from the ‘nnet’ package and likelihood-ratio tests with *Anova()* from ‘car’.

### Ethics

ATLAS research protocol (version 3.0, October 8 2020) has been approved by the WHO Ethical Research Committee (January 12, 2021, reference: ERC 0003181), the National Ethics Committee for Life Sciences and Health of Côte d’Ivoire (November 27, 2020, reference: 191-20/MSHP/CNESVS-km, IRB:000111917), the Ethics Committee of the Faculty of Medicine and Pharmacy of the University of Bamako, Mali (November 16, 2020, reference: 2020/254/CE/FMPOS/FAPH), and the National Ethics Committee for Health Research of Senegal (January 26, 2021, protocol SEN19/32, n°8 MSAS/CNERS/Sec).

The full research protocol was written in French (https://hal.science/ATLAS_ADVIH/hal-04121482v1). The peer-reviewed protocol has been published in English elsewhere [30].

## Results

### HIVST results

Of the 2 615 participants recruited in phase 1, 2 346 (89.7%) reported a self-interpreted HIVST result consistent with their reported number of visibles lines on the HIVST: 2 292 (88.0%) reported one line self-interpreted as non-reactive, 50 (1.9%) two lines self-interpreted as reactive, and 4 (0.2%) no/one line self-interpreted as invalid (table 1). In contrast, 48 (1.8%) reported an inconsistent response: 10 (0.4%) one line self-interpreted as reactive, 35 (1.3%) two lines self-interpreted as non-reactive/ and 3 (0.1%) no line self-interpreted as non-reactive. Finally, 221 (8.5%) reported a partial result: 147 (5.6%) reported 0, 1 or 2 lines but did not know how to interpret the result or refused to answer, 46 (1.7%) self-interpreted their result but did not know or refused to report the number of lines, and 28 (1.1%) did not know or refused to answer to both questions.

**Table 1.**
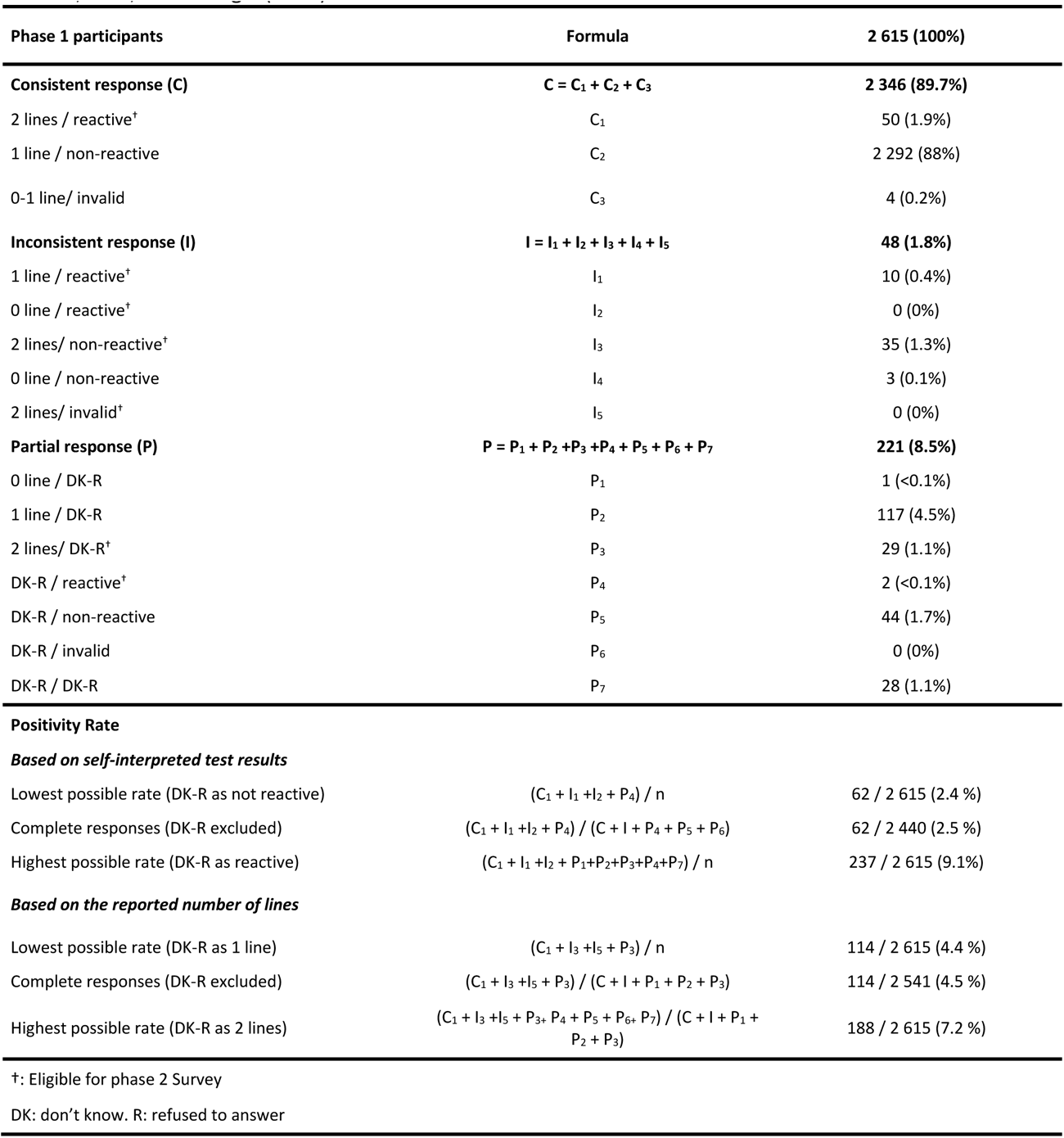
Reported self-interpreted HIV self-test (HIVST) result, reported number of lines on the HIVST, and positivity rates according to different hypotheses among participants of the first phase of the survey in Côte d’Ivoire, Mali, and Senegal (2021).

**Table 2.** Linkage to confirmatory testing, proportion being confirmed HIV positive and treatment initiation, by reported number of lines and self-interpreted HIVST result among eligible participants of the second phase of the survey who completed their questionnaire in Côte d’Ivoire, Mali, and Senegal (2021).

| Reported number of lines/<br>self-interpreted HIVST result | Completed<br>phase 2 | Linked to confirmatory testing |  | Confirmed HIV positive |  | Initiated ART |  |
| --- | --- | --- | --- | --- | --- | --- | --- |
|  | n | n (%) | 95%CI | n (%) | 95%CI | n (%) | 95%CI |
| Overall | 78 | 34 (44%) | 33% to 55% | 19 (56%) | 38% to 72% | 18 (95%) | 72% to 100% |
| 2 lines / reactive | 27 | 15 (56%) | 36% to 74% | 12 (80%) | 51% to 95% | 12 (100%) | 70% to 100% |
| 1 line / reactive | 7 | 1 (14%) | 1% to 58% | 0 (0%) | 0% to 80% |  |  |
| 2 lines / non-reactive | 25 | 9 (36%) | 19% to 57% | 3 (33%) | 9% to 69% | 3 (100%) | 31% to 100% |
| 2 lines / DK-R | 18 | 8 (44%) | 22% to 69% | 4 (50%) | 22% to 78% | 3 (75%) | 22% to 99% |
| DK-R / reactive | 1 | 1(100%) | 5% to 100% | 0 (0%) | 0% to 95% |  |  |

DK: don't know. R: refuse to answer. CI: confidence interval.

### HIVST positivity rates

Based on the self-interpreted HIVST results, the overall positivity rate was 2.5% when only complete responses were considered (Table 1). It would have been similar (2.4%) if DK-R responses were considered non-reactive (lowest possible rate). Considering DK-Rs as reactive would have increased the positivity rate to 9.1% (highest possible rate). Based on the estimated number of visible lines, the overall positivity rate was 4.5% (complete responses, lowest possible rate: 4.4%, highest possible rate: 7.2%).

Multivariable models did not show any significant effect of key population profile, country, age group, marital status, or being a first-time tester on positivity rates (Tables S1a and S1b). No effect of educational level was observed on positivity rates based on the reported number of visible lines. However, a significant effect of the educational level was observed on positivity rates based on self-reported HIVST results (p=0.002, q=0.014): individuals with a secondary or a higher level of education have a higher probability of reporting a reactive test (adjusted OR equal to 4.00 [95% confidence interval: 1.44 to 12.9] and 4.12 [1.76 to 12.1] respectively).

Although not statistically significant, we observed variations between key population profiles (Figure 3, Table S2). Based on self-reported results, positivity rates were 3.4% for men [possible range from 3.2 to 9.8%] and 1.0% for women [1.0 to 2.9%] in MSM-based channels, 1.7% for men [1.6 to 8.2%] and 2.7% [2.5 to 10.0%] for women in FSW-based channels, vs 0.8% for men [0.7 to 5.8%] and 1.5% for women [1.4 to 8.2%] in the other distribution channels (PWUD-based channels, index testing and STI consultations). Observed positivity rates varied by age group (Table S3): 2.4% for 15-24 years old [2.2 to 7.8%], compared to 2.9% for 25-34 years old [2.7 to 9.5%] and 2.0% for those aged 35 years or older [1.8 to 12.0%].

**Figure 3.**
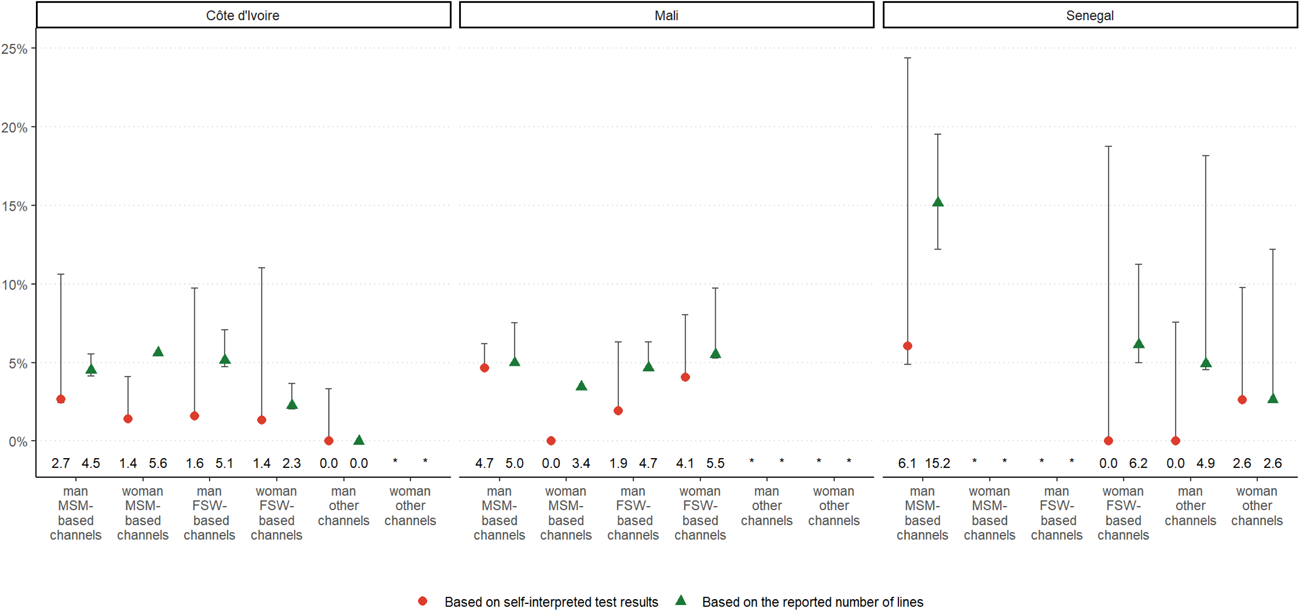
Positivity rates based on self-interpreted HIVST results or the reported number of visible lines, by key population profiles and country, among participants of the first survey phase in Côte d’Ivoire, Mali, and Senegal (2021). Error bars indicate possible range. An asterisk indicates that there was less than 25 participants in that distribution channel. FSW=female sex worker, MSM=men who have sex with men. MSM-based channels include facility-based and outreach. FSW-based channels include facility-based and outreach. Other channels include PWUD-based channels, index testing and STI consultations.

### Participation in phase 2

During phase 1, 126 individuals reported two lines or self-interpreted their result as reactive and were therefore eligible for phase 2 (Table 1). Among them, 6 refused to be recontacted after phase 1 (Figure 4). Among the 120 (95%) who agreed to be recontacted, 24 (20%) were unreachable at the time of the phase 2 survey, and 96 (80%) were successfully recontacted. Among the latest, 89 (93%) accepted to participate in the phase 2 survey. Ten dropped out before the end of the interview, and 1 disconnected and was unreachable afterwards. As a result, 78 participants completed phase 2 questionnaire. Of the 78 participants, 39 (50%) were from Côte d’Ivoire, 31 (40%) from Mali, and 8 (10%) from Senegal (Table S2). Participation rates were 54% (27/50) for participants who reported a consistent response (2 lines and reactive), 71% (32/45) for those with an inconsistent response (either 2 lines & non-reactive, or 1 line & reactive), and 66% (19/31) for those reporting a partial response (2 lines & DK-R or DK-R & reactive).

**Figure 4.**
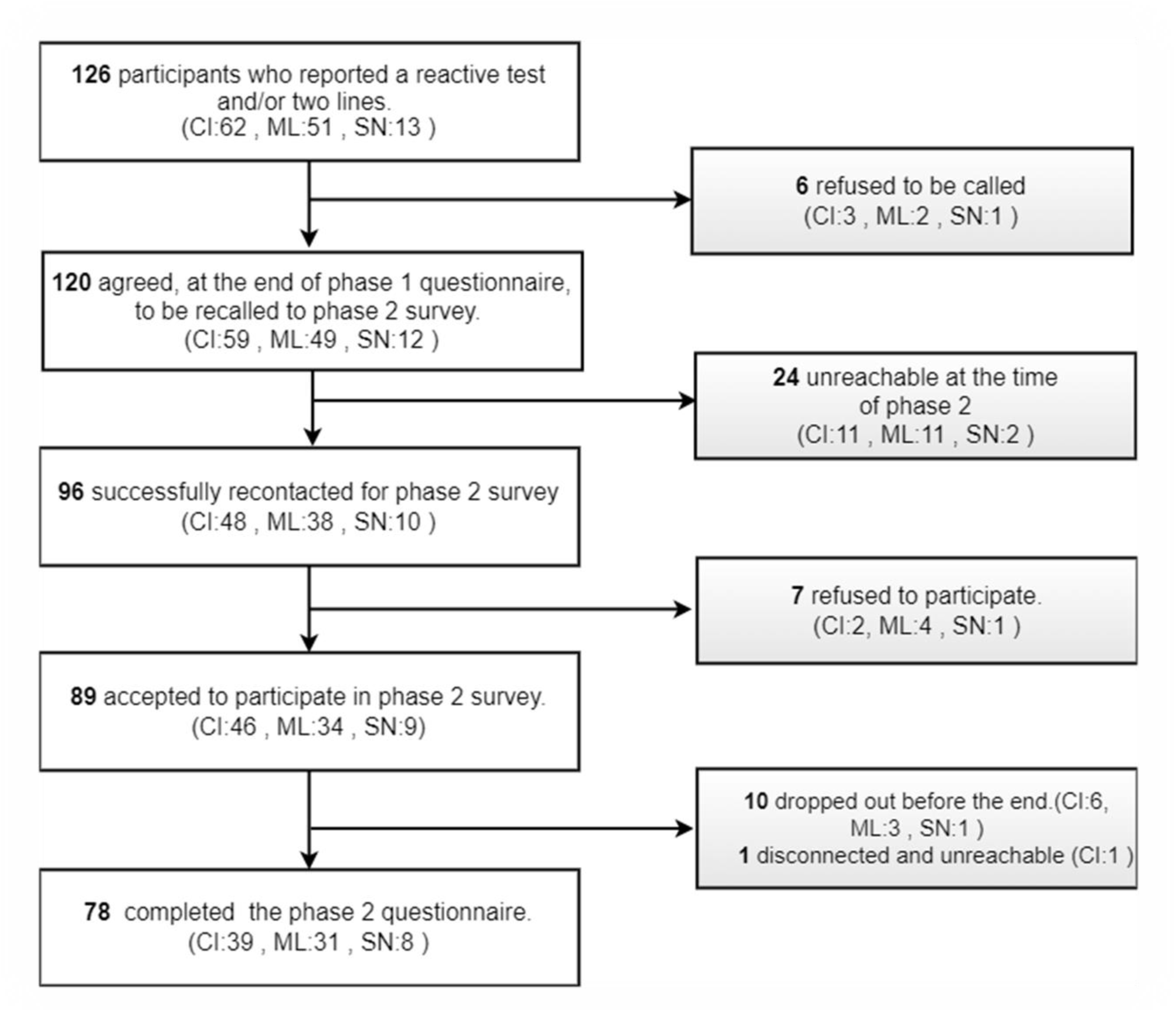
Flow chart of the participant selection process for the 2nd phase of the survey in Côte d’Ivoire (CI), Mali (ML), and Senegal (SN) (2021).

The participants who completed the phase 2 questionnaire had similar sociodemographic characteristics (e.g. country, sex, distribution channel, age group, marital status) compared to those eligible for phase 2, but that did not complete it, and to phase 1 participants not eligible for phase 2 (Table S4). For most participants (86%), phase 2 questionnaire was completed between 4 and 6 months after phase 1 questionnaire (Table S5).

### Linkage to confirmatory testing and care

Overall, 34 of the 78 who completed the phase 2 questionnaire (44%) reported having performed confirmatory testing. Linkage was higher for those who reported 2 lines and correctly self-interpreted their result as reactive (56%,95%CI: 36-74%), than for those who reported two lines but did not know or refused to report their test interpretation (44%, (95%CI: 22-69%) and those who reported 2 lines but incorrectly self-interpreted the result as non-reactive (36%, 95%CI: 19-57%) (Table 3). Finally, among the 8 participants who reported none/one line or did not know how many lines and incorrectly self-interpreted the result as reactive, only 2 linked to confirmatory testing.

The main reason given for not linking to confirmatory testing was that “*their HIVST was non-reactive*” (18/44, 41%, and although 8 of these 18 reported a reactive result in phase 1 questionnaire), followed by “*not knowing that a confirmation test was required*” (10/44, 23%), and “*not having time*” (8/44, 18%) (Table S6).

When participants were linked to confirmatory testing, it was usually shortly after performing their HIVST: 53% linked in less than one week and 91% in less than 3 months (Table S5). Most participants (65%) performed their confirmatory testing in a general public facility (health centre, hospital, clinic or maternity) wheras 35% chose a community-based clinic or health centre dedicated to key populations (Table S7).

Among the 34 that linked to confirmatory testing, 19 (56%, 95%CI: 38-72%) were confirmed HIV-positive, and 18 (95%, 95%CI; 72-100%) initiated antiretroviral treatment. Of the 18 participants who initiated ART, 11 (72%) underwent their confirmation test less than a week after their self-test, 2 (11%) did so between 1 and 2 weeks, 1 (5.6%) between 3 and 4 weeks, 1 (5.6%) waited between 1 and 2 months, and 1 (5.6%) proceeded with the test three months later. Among the 27 who reported a consistent reactive response in the phase 1 questionnaire, 15 (56%, 95%CI: 36-74%) linked to confirmatory test, 12 (80%) were confirmed HIV-positive and all started treatment (100%).

## Discussion

Our study shows that the strategy implemented by the ATLAS program, through primary and secondary distribution of HIVST kits and dedicated channels, achieved HIV positivity rates of 2.5% (central hypothesis, low: 2.4%, high: 9.1%) based on self-interpreted results, and 4.5% (central hypothesis, low: 4.4%, high: 7.2%) based on the reported number of lines. The proportion of participants with a reactive HIVST that sought confirmatory testing was 44% (95% CI: 33%-55%). Of those who underwent confirmatory testing, 56% (95% CI: 38%-72%) were found to be HIV-positive and, among them, 95% (95% CI: 72%-100%) initiated treatment. Among the participants who confirmed their reactive HIVST with a traditional facility-based HIV test, 65% did so within a week and 91% within three months.

According to our estimates, HIVST positivity rates in Côte d’Ivoire were 2.0% (complete responses, lowest possible: 1.8%, highest possible: 9.8%) based on self-interpreted results and 3.9% (3.8% to 5.4%) based on the number of lines reported. In Mali, these rates were respectively 3.6% (3.5 to 6.7%) and 5.0% (4.9% to 7.8%), while, in Senegal, they were 1.4% (1.2 to 15.0%) and 6.0% (5.4% to 14.9%). Overall, these results for HIVST positivity are generally higher than the average overall positivity of HIV testing services (excluding HIVST) in West Africa. For instance, in 2020 an estimated 1.9% of all HIV tests performed were found to be positive in the region (95% credible intervals: 1.3 to 2.7%) [42]. Our results are in line with data collected by ATLAS implementing partners. Between 2020 and 2021, these ATLAS partners collected spontaneous feedback from HIVST users. This unpublished data collection was non-systematic and varied from one partner to another. Among 4 463 documented feedbacks, HIVST was reactive for 188 cases (4.2%), consistent with our estimates based on the reported number of visible lines (4.5%). In 2021, a study based on the UNAIDS-supported *Shiny90* mathematical model [43] estimated, using data from 184 population surveys and reports from national HIV screening programs from 40 sub-Saharan African countries, that the positivity rates for conventional HIV testing were 1.4% in Côte d’Ivoire, 2.2% in Mali, and 1.0% in Senegal. Our estimates for HIVST were higher than these estimates for convential testing. Collectively, these results provide evidence that HIVST is a high-yield testing modality that can address the unmet HIV testing needs of key populations and their partners.

It is important to interpret HIV positivity rates while considering the treatment-adjusted prevalence (i.e., removing those on treatment from the numerator and denominator of HIV prevalence), a more reliable indicator for evaluating the effectiveness and positivity rates of targeted screening programs [44]. In West Africa, the treatment-adjusted prevalence remained relatively low in 2021: 0.6% in Côte d’Ivoire, 0.7% in Mali, and 0.06% in Senegal, according to UNAIDS data (https://aidsinfo.unaids.org/). Our positivy rates in each country are higher than the treatment-adjusted prevalence, suggesting that the ATLAS HIVST distribution strategy successfully reached a hard-to-reach population and at positivity levels at least as high as with passive surveillance.

In our study, 2.0% of the participants reported an inconsistent response between the number of visible lines and their self-interpretation of the result and 6.0% reported a number of lines but didn’t know how to interpret it or refused to answer, suggesting potential issues in interpreting the number of visible lines on HIVST kits. In the context of the ATLAS program, the distribution strategy combining primary and secondary approaches has led many HIVST users to perform their HIVST without receiving advice from a healthcare professional or a trained peer educator. Although the HIVST is not designed to require supervision, it is essential to have received information on its use before proceeding with the test. A study conducted within the framework of the ATLAS program demonstrated that the manufacturer’s instructions alone were insufficient in a multilingual context with low literacy levels. The use of additional aid, such as a demonstration video or a toll-free helpline, proved to be necessary [45]. Similarly, a study carried out in China in 2018 on the unsupervised use of HIVST among 27 MSM found that only 5 (or 19%) made no errors, and 44% received an invalid test result due to various mistakes made [46]. However, the lack of supervision is likely insufficient to explain the inconsistencies observed [23]. Some inconsistencies may result from a misunderstanding of the terms “*reactive*” and “*non-reactive*”, particularly considering that HIVST was a new tool in our context and that traditional terms used to describe conventional HIV testing are “*positive*” and “*negative*”. This possible misunderstanding of the terms is also highlighted by the fact that 8 participants reported a “*reactive*” result in phase 1 questionnaire and then in phase 2 that their test was “*non-reactive*” as the main reason for not linking to confirmatory testing. It is also suggested by the fact that, in our multivariable logistic regression models, individuals with a low level of education were significantly less likely to report a reactive HIVST result, while no significant difference was observed regarding the reported number of visible lines. Specific qualitative interviews or focus groups discussion with HIVST users could help better understand how they perceive different terms.

Linkage to confirmatory testing following a reactive test was 44% (95% confidence interval from 33% to 55%). However, this estimate includes some individuals who did not adequately self-interpreted their HIVST result as reactive. When considering only those who reported two lines and self-interpreted their result as reactive, the linkage rate increased to 56% (36% to 74%). This percentage is closer to that was observed in a study conducted in Kenya on HIV testing of FSW male partners using HIVST secondary distribution, where 65% of men with a reactive result had a confirmatory test [47]. Our estimates were based on small numbers resulting in large confidence intervals, but are still showing a low rate.

Linkage to confirmatory testing happened relatively quickly after HIVST use: 53% did it in less than a week and 91% in less than three months. Similar results were observed in a study in the general population in Zambia[48], and a study among MSM in Nigeria [49].

The main reasons given for not linking to confirmatory testing suggest potential misinterpretation of the result or misunderstanding about the need to perform a confirmatory HIV test, highlighting the need to improve messaging around HIVST, in particular when HIV self-testing policies will be scaled-up. For those who did confirmatory testing and were confirmed HIV positive, initiation of antiretroviral treatment was almost systematic, showing good linkage to care after confirmatory testing, as observed in many HIVST studies in sub-Saharan Africa [50–52].

Previous analyses of ATLAS data showed that HIVST could reach people not reached by conventional HIV testing approaches [53], particularly partners and clients of key populations and key population members not self-identifying as such [54]. It is consistent with the finding that two-thirds of participants who did confirmatory testing went to a general health facility rather than a community clinic dedicated to key populations. In a study conducted in 2018 in Côte d’Ivoire among MSM, one-third of the participants preferred community-based testing, one-third expressed no preference, and one-third preferred undifferentiated HIV testing services (general population), mentioning the lack of discretion and anonymity of community-based sites and the desire to avoid the gaze of others [55].

The implementation of a telephone survey, aimed at gathering information from HIVST users while preserving anonymity and without interfering with secondary distribution, has proven to be very useful to evaluate the ATLAS program. However, its high cost makes it difficult to integrate it into national strategies for assessing the impact of HIVST. In addition, due to the small number of observations, we had low statistical power regarding the estimates of positivity rates and linkage to confirmatory testing. Nevertheless, other impact evaluation methods, such as data triangulation [36] and modelling [37], may prove more suitable for routine monitoring of HIVST’s impacts.

A previous analysis of this survey among ATLAS HIVST users showed that HIVST secondary distribution was feasible and acceptable [39]: participants reported that they appreciated the ease of use of HIVST, its discretion and the fact that they are autonomous in carrying out the test. Finally, HIVST appeared as a relevant additional approach for those usually distant from community activities and HIV testing services, and has the potential to reach, beyond key populations, partners, clients, and other groups vulnerable to HIV.

ATLAS’ HIVST distribution strategy successfully reached people living with HIV in West Africa, although linkage to confirmatory testing following a reactive HIVST remained relatively low in these first years of HIVST implementation, and sub-optimal in the perspective of achieving UNAIDS 95-95-95 targets. However, among participants who confirmed their reactive self-test result with a traditional facility-based HIV test, a substantial proportion quickly proceeded with this confirmation (more than half in less than a week and the vast majority in less than three months). Furthermore, if individuals were confirmed HIV-positive, almost all began antiretroviral treatment. We showed that HIVST has the potential to reach more hidden populations and constitutes a relevant complementary tool to existing screening services. To fully harness the potential of self-tests, messaging around HIVST and its interpretation could be improved.

## Data Availability

Data, scripts and code are available online on the Zenodo website.

https://doi.org/10.5281/zenodo.11061878

https://doi.org/10.5281/zenodo.10210464

## Acknowledgements

We wish to acknowledge the commitment and determination of all the ATLAS program teams, which made this research possible. We would also like to thank the interviewers for their professionalism in collecting this sensitive data. Finally, we are grateful to the participants who were kind enough to give us some of their time by agreeing to take part in the survey.

## Data, scripts, code, and supplementary information availability

Data, scripts and code are available online (https://doi.org/10.5281/zenodo.11086135) as well as the survey questionnaires (https://doi.org/10.5281/zenodo.10210464). Supplementary figures and tables are provided in the appendices.

## Appendices

**Table S1a:**
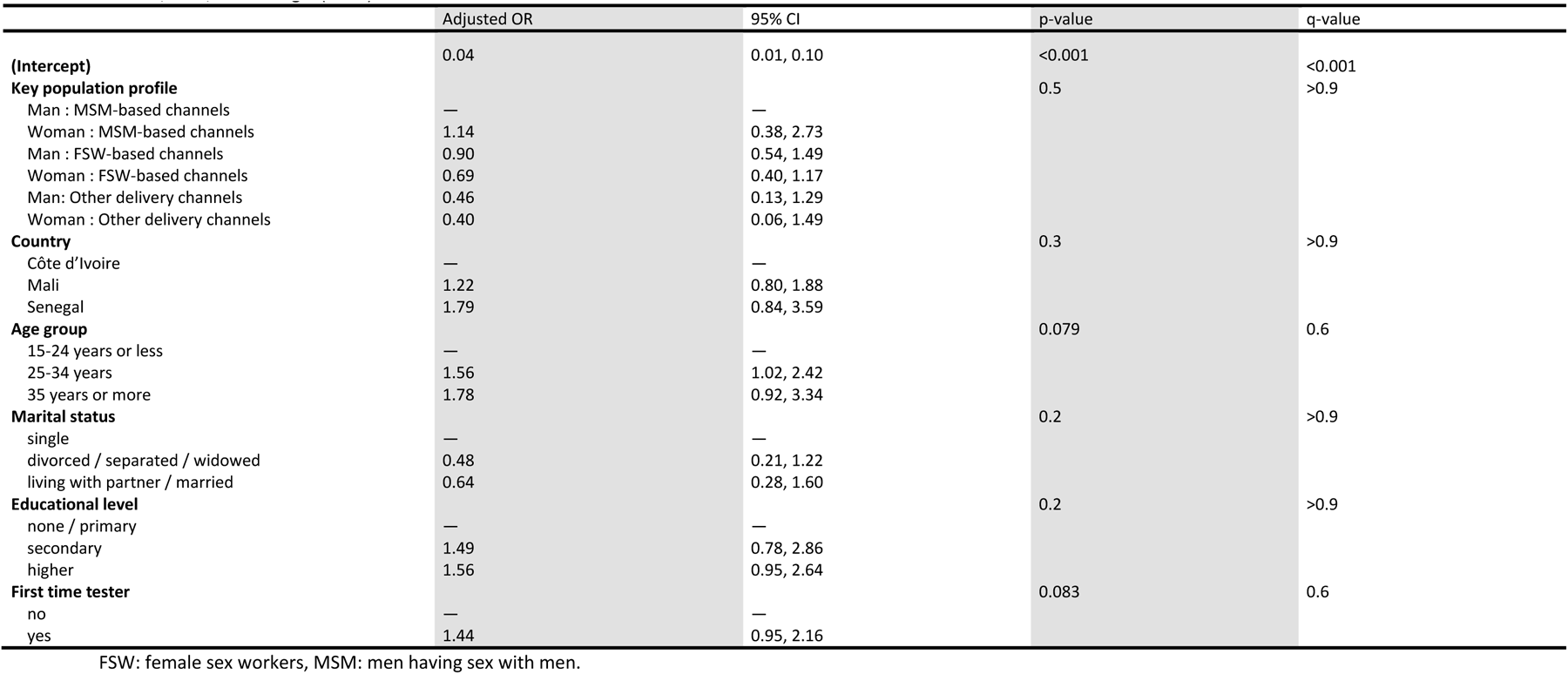
Factors associated (logistic regression) with positivity rate based on the reported number of visible lines among participants of the first survey phase in Côte d’Ivoire, Mali, and Senegal (2021)

|  | Adjusted OR | 95% CI | p-value | q-value |
| --- | --- | --- | --- | --- |
| **(Intercept)** | 0.04 | 0.01, 0.10 | <0.001 | <0.001 |
| **Key population profile** |  |  | 0.5 | >0.9 |
| Man : MSM-based channels | — | — |  |  |
| Woman : MSM-based channels | 1.14 | 0.38, 2.73 |  |  |
| Man : FSW-based channels | 0.90 | 0.54, 1.49 |  |  |
| Woman : FSW-based channels | 0.69 | 0.40, 1.17 |  |  |
| Man: Other delivery channels | 0.46 | 0.13, 1.29 |  |  |
| Woman : Other delivery channels | 0.40 | 0.06, 1.49 |  |  |
| **Country** |  |  | 0.3 | >0.9 |
| Côte d'Ivoire | — | — |  |  |
| Mali | 1.22 | 0.80, 1.88 |  |  |
| Senegal | 1.79 | 0.84, 3.59 |  |  |
| **Age group** |  |  | 0.079 | 0.6 |
| 15-24 years or less | — | — |  |  |
| 25-34 years | 1.56 | 1.02, 2.42 |  |  |
| 35 years or more | 1.78 | 0.92, 3.34 |  |  |
| **Marital status** |  |  | 0.2 | >0.9 |
| single | — | — |  |  |
| divorced / separated / widowed | 0.48 | 0.21, 1.22 |  |  |
| living with partner / married | 0.64 | 0.28, 1.60 |  |  |
| **Educational level** |  |  | 0.2 | >0.9 |
| none / primary | — | — |  |  |
| secondary | 1.49 | 0.78, 2.86 |  |  |
| higher | 1.56 | 0.95, 2.64 |  |  |
| **First time tester** |  |  | 0.083 | 0.6 |
| no | — | — |  |  |
| yes | 1.44 | 0.95, 2.16 |  |  |

FSW: female sex workers, MSM: men having sex with men.

**Table S1b:**
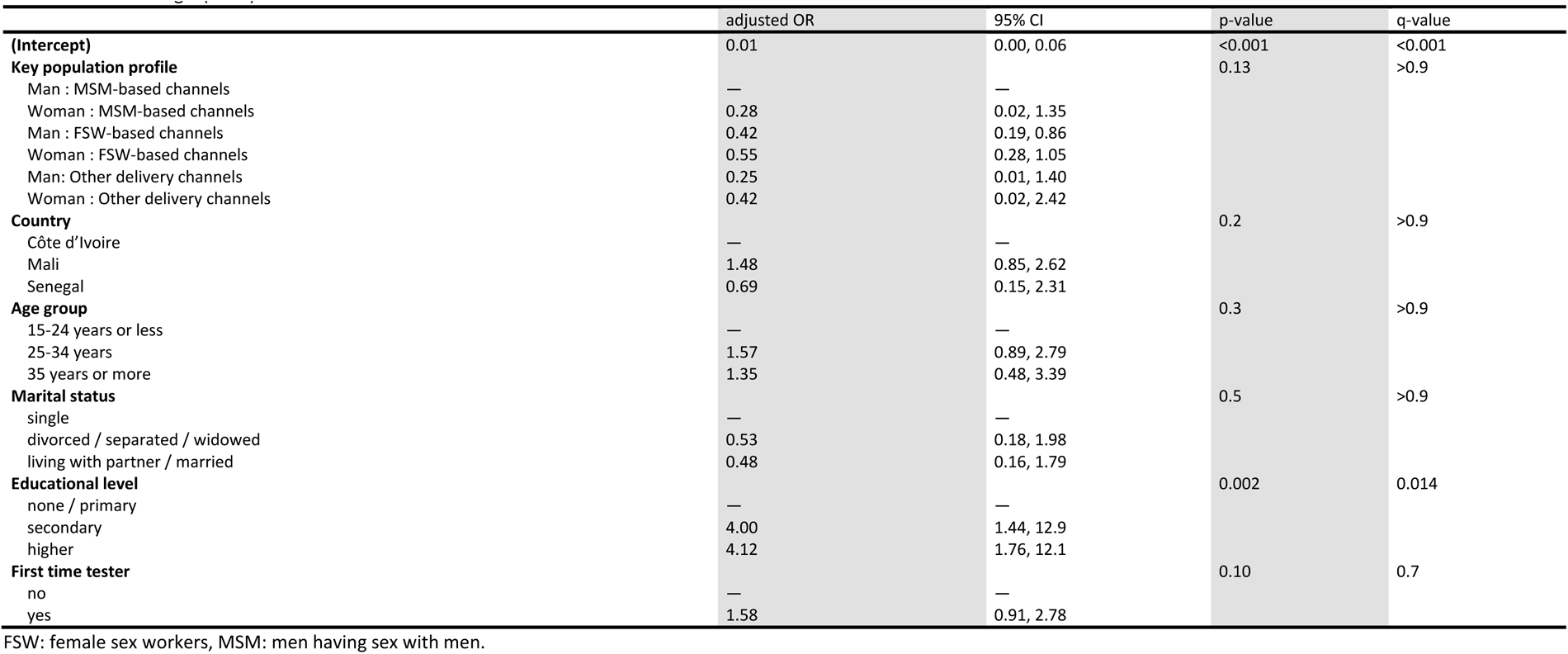
Factors associated (logistic regression) with positivity rate based on self-reported HIVST, among participants of the first survey phase in Côte d’Ivoire, Mali, and Senegal (2021)

|  | adjusted OR | 95% CI | p-value | q-value |
| --- | --- | --- | --- | --- |
| **(Intercept)** | 0.01 | 0.00, 0.06 | <0.001 | <0.001 |
| **Key population profile** |  |  | 0.13 | >0.9 |
| Man : MSM-based channels | — | — |  |  |
| Woman : MSM-based channels | 0.28 | 0.02, 1.35 |  |  |
| Man : FSW-based channels | 0.42 | 0.19, 0.86 |  |  |
| Woman : FSW-based channels | 0.55 | 0.28, 1.05 |  |  |
| Man: Other delivery channels | 0.25 | 0.01, 1.40 |  |  |
| Woman : Other delivery channels | 0.42 | 0.02, 2.42 |  |  |
| **Country** |  |  | 0.2 | >0.9 |
| Côte d'Ivoire | — | — |  |  |
| Mali | 1.48 | 0.85, 2.62 |  |  |
| Senegal | 0.69 | 0.15, 2.31 |  |  |
| **Age group** |  |  | 0.3 | >0.9 |
| 15-24 years or less | — | — |  |  |
| 25-34 years | 1.57 | 0.89, 2.79 |  |  |
| 35 years or more | 1.35 | 0.48, 3.39 |  |  |
| **Marital status** |  |  | 0.5 | >0.9 |
| single | — | — |  |  |
| divorced / separated / widowed | 0.53 | 0.18, 1.98 |  |  |
| living with partner / married | 0.48 | 0.16, 1.79 |  |  |
| **Educational level** |  |  | 0.002 | 0.014 |
| none / primary | — | — |  |  |
| secondary | 4.00 | 1.44, 12.9 |  |  |
| higher | 4.12 | 1.76, 12.1 |  |  |
| **First time tester** |  |  | 0.10 | 0.7 |
| no | — | — |  |  |
| yes | 1.58 | 0.91, 2.78 |  |  |

FSW: female sex workers, MSM: men having sex with men.

**Table S2.** Positivity rates based on self-interpreted HIVST results or the reported number of visible lines, by distribution channel, sex and country, among participants of the first survey phase in Côte d’Ivoire, Mali, and Senegal (2021). FSW-based channels include facility-based and outreach. Other channels include PWUD-based channels, index testing and STI consultations.

|  |  |  | MSM-based channels |  | FSW-based channels |  | Others delivery channels |  | Total |
| --- | --- | --- | --- | --- | --- | --- | --- | --- | --- |
|  |  |  | Man | Woman | Man | Woman | Man | Woman |  |
| Positivity rate based on self-reported HIVST results | Lowest possible rate | Côte d'Ivoire | 2.5% (16/650) | 1.4% (1/73) | 1.5% (5/339) | 1.2% (3/245) | 0% (0/60) | 0% (0/23) † | 1.8% (25/1 390) |
|  |  | Mali | 4.6% (14/306) | 0% (0/29) | 1.9% (5/269) | 3.9% (14/360) | 9.1% (1/11) † | 0% (0/9) † | 3.5% (34/984) |
|  |  | Senegal | 4.9% (2/41) | 0% (0/1) † | 0% (0/12) † | 0% (0/80) | 0% (0/66) | 2.4% (1/41) | 1.2% (3/241) |
|  |  | Overall | 3.2% (32/997) | 1.0% (1/103) | 1.6% (10/620) | 2.5% (17/685) | 0.7% (1/137) | 1.4% (1/73) | 2.4% (62/2 615) |
|  | Complete responses | Côte d'Ivoire | 2.7% (16/597) | 1.4% (1/71) | 1.6% (5/311) | 1.4% (3/221) | 0% (0/58) | 0% (0/21) † | 2.0% (25/1 279) |
|  |  | Mali | 4.7% (14/301) | 0% (0/29) | 1.9% (5/257) | 4.1% (14/345) | 9.1% (1/11) † | 0% (0/9) † | 3.6% (34/952) |
|  |  | Senegal | 6.1% (2/33) | 0% (0/1) † | 0% (0/11) † | 0% (0/65) | 0% (0/61) | 2.6% (1/38) | 1.4% (3/209) |
|  |  | Overall | 3.4% (32/931) | 1.0% (1/101) | 1.7% (10/579) | 2.7% (17/631) | 0.8% (1/130) | 1.5% (1/68) | 2.5% (62/2 440) |
|  | Highest possible rate | Côte d'Ivoire | 10.6% (69/650) | 4.1% (3/73) | 9.7% (33/339) | 11% (27/245) | 3.3% (2/60) | 8.7% (2/23) | 9.8% (136/1 390) |
|  |  | Mali | 6.2% (19/306) | 0% (0/29) | 6.3% (17/269) | 8.1% (29/360) | 9.1% (1/11) † | 0% (0/9) † | 6.7% (66/984) |
|  |  | Senegal | 24.0% (10/41) | 0.0% (0/1) † | 8.3% (1/12) † | 19.0% (15/80) | 7.6% (5/66) | 9.8% (4/41) | 15.0% (35/241) |
|  |  | Overall | 9.8% (98/997) | 2.9% (3/103) | 8.2% (51/620) | 10.0% (71/685) | 5.8% (8/137) | 8.2% (6/73) | 9.1% (237/2 615) |
| Positivity rate based on the reported number of visible lines | Lowest possible rate | Côte d'Ivoire | 4.2% (27/650) | 5.5% (4/73) | 4.7% (16/339) | 2.0% (5/245) | 0% (0/60) | 4.3% (1/23) | 3.8% (53/1 390) |
|  |  | Mali | 4.9% (15/306) | 3.4% (1/29) | 4.5% (12/269) | 5.3% (19/360) | 9.1% (1/11) † | 0% (0/9) † | 4.9% (48/984) |
|  |  | Senegal | 12.2% (5/41) | 0% (0/1) † | 0% (0/12) † | 5.0% (4/80) | 4.5% (3/66) | 2.4% (1/41) | 5.4% (13/241) |
|  |  | Overall | 4.7% (47/997) | 4.9% (5/103) | 4.5% (28/620) | 4.1% (28/685) | 2.9% (4/137) | 2.7% (2/73) | 4.4% (114/2 615) |
|  | Complete responses | Côte d'Ivoire | 4.2% (27/641) | 5.5% (4/73) | 4.8% (16/331) | 2.1% (5/241) | 0% (0/60) | 4.5% (1/22) † | 3.9% (53/1 368) |
|  |  | Mali | 5.0% (15/298) | 3.4% (1/29) | 4.5% (12/264) | 5.5% (19/344) | 9.1% (1/11) † | 0% (0/9) † | 5.0% (48/955) |
|  |  | Senegal | 13.2% (5/38) | 0% (0/1) † | 0% (0/10) † | 5.3% (4/75) | 5.3% (3/57) | 2.7% (1/37) | 6.0% (13/218) |
|  |  | Overall | 4.8% (47/977) | 4.9% (5/103) | 4.6% (28/605) | 4.2% (28/660) | 3.1% (4/128) | 2.9% (2/68) | 4.5% (114/2 541) |
|  | Highest possible rate | Côte d'Ivoire | 5.5% (36/650) | 5.5% (4/73) | 7.1% (24/339) | 3.7% (9/245) | 0% (0/60) | 8.7% (2/23) † | 5.4% (75/1 390) |
|  |  | Mali | 7.5% (23/306) | 3.4% (1/29) | 6.3% (17/269) | 9.7% (35/360) | 9.1% (1/11) † | 0% (0/9) † | 7.8% (77/984) |
|  |  | Senegal | 19.5% (8/41) | 0% (0/1) † | 16.7% (2/12) † | 11.2% (9/80) | 18.2% (12/66) | 12.2% (5/41) | 14.9% (36/241) |
|  |  | Overall | 6.7% (67/997) | 4.9% (5/103) | 6.9% (43/620) | 7.7% (53/685) | 9.5% (13/137) | 9.6% (7/73) | 7.2% (188/2 615) |

DK: don't know. R: refusals. FSW: female sex workers, MSM: men having sex with men, PR: positivity rate.
†: indicated cells with less than 25 participants.
Lowest possible rate: DK-R as non-reactive or 1 line. Complete responses: DK-R excluded from the numerator and the denominator. Highest possible rate: DK-R as reactive or 2 lines.

**Table S3.** Positivity rates based on self-interpreted HIVST results or the reported number of visible lines, by age group and country, among participants of the first survey phase in Côte d’Ivoire, Mali, and Senegal (2021).

|  |  |  | 15-24 years | 25-34 years old | 35 years or more | Total |
| --- | --- | --- | --- | --- | --- | --- |
| Positivity rate based on self-reported HIVST results | Lowest possible rate | Côte d'Ivoire | 1.7% (11/645) | 2.0% (11/553) | 1.6% (3/192) | 1.8% (25/1 390) |
|  |  | Mali | 3.3% (15/455) | 3.9% (16/415) | 2.6% (3/114) | 3.5% (34/984) |
|  |  | Senegal | 0.0% (0/64) | 2.1% (2/95) | 1.2% (1/82) | 1.2% (3/241) |
|  |  | Overall | 2.2% (26/1 164) | 2.7% (29/1 063) | 1.8% (7/388) | 2.4% (62/2 615) |
|  | Complete responses | Côte d'Ivoire | 1.8% (11/604) | 2.2% (11/506) | 1.8% (3/169) | 2.0% (25/1 279) |
|  |  | Mali | 3.4% (15/439) | 4.0% (16/403) | 2.7% (3/110) | 3.6% (34/952) |
|  |  | Senegal | 0.0% (0/56) | 2.4% (2/82) | 1.4% (1/71) | 1.4% (3/209) |
|  |  | Overall | 2.4% (26/1 099) | 2.9% (29/991) | 2.0% (7/350) | 2.5% (62/2 440) |
|  | Highest possible rate | Côte d'Ivoire | 8.1% (52/645) | 10.0% (58/553) | 14.0% (26/192) | 9.8% (136/1 390) |
|  |  | Mali | 6.8% (31/455) | 6.7% (28/415) | 6.1% (7/114) | 6.7% (66/984) |
|  |  | Senegal | 13.0% (8/64) | 16.0% (15/95) | 15.0% (12/82) | 15.0% (35/241) |
|  |  | Overall | 7.8% (91/1 164) | 9.5% (101/1 063) | 12.0% (45/388) | 9.1% (237/2 615) |
| Positivity rate based on the reported number of visible lines | Lowest possible rate | Côte d'Ivoire | 3.1% (20/645) | 4.5% (25/553) | 4.2% (8/192) | 3.8% (53/1 390) |
|  |  | Mali | 4.8% (22/455) | 4.8% (20/415) | 5.3% (6/114) | 4.9% (48/984) |
|  |  | Senegal | 1.6% (1/64) | 7.4% (7/95) | 6.1% (5/82) | 5.4% (13/241) |
|  |  | Overall | 3.7% (43/1 164) | 4.9% (52/1 063) | 4.9% (19/388) | 4.4% (114/2 615) |
|  | Complete responses | Côte d'Ivoire | 3.1% (20/637) | 4.6% (25/546) | 4.3% (8/185) | 3.9% (53/1 368) |
|  |  | Mali | 4.9% (22/447) | 5.0% (20/401) | 5.6% (6/107) | 5.0% (48/955) |
|  |  | Senegal | 1.9% (1/54) | 8.2% (7/85) | 6.3% (5/79) | 6.0% (13/218) |
|  |  | Overall | 3.8% (43/1 138) | 5.0% (52/1 032) | 5.1% (19/371) | 4.5% (114/2 541) |
|  | Highest possible rate | Côte d'Ivoire | 4.3% (28/645) | 5.8% (32/553) | 7.8% (15/192) | 5.4% (75/1 390) |
|  |  | Mali | 6.6% (30/455) | 8.2% (34/415) | 11.0% (13/114) | 7.8% (77/984) |
|  |  | Senegal | 17.0% (11/64) | 18.0% (17/95) | 9.8% (8/82) | 15.0% (36/241) |
|  |  | Overall | 5.9% (69/1 164) | 7.8% (83/1 063) | 9.3% (36/388) | 7.2% (188/2 615) |

**Table S4.** Eligibility and participation in phase 2 survey by sociodemographic characteristics, distribution channel, and HIV testing history (bivariable comparison and multivariable multinomial regression model). FSW-based channels and MSM-based channels include facility-based and outreach. Other channels include PWUD-based channels, index testing and STI consultations.

|  | completed phase 2<br>questionnaire<br>N = 78 | eligible for phase 2 but did not<br>complete the questionnaire<br>N = 48 | phase 1 participants not<br>eligible for phase 2<br>N = 2 489 | Bivariable<br>comparison<br>p-value<br>(Chi<sup>2</sup> test) | Multivariable multinomial<br>regression model<br>p-value | Overall<br>N = 2 615<br>(phase 1<br>participants) |
| --- | --- | --- | --- | --- | --- | --- |
| **Country** |  |  |  | 0.9 | 0.8 |  |
| Côte d'Ivoire | 39 (50%) | 23 (48%) | 1 328 (53%) |  |  | 1 390 (53%) |
| Mali | 31 (40%) | 20 (42%) | 933 (37%) |  |  | 984 (38%) |
| Senegal | 8 (10%) | 5 (10%) | 228 (9.2%) |  |  | 241 (9.2%) |
| **Sex and distribution channel** |  |  |  | 0.3 | 0.06 |  |
| Man: MSM-based channels | 35 (45%) | 18 (38%) | 944 (38%) |  |  | 997 (38%) |
| Woman: MSM-based channels | 5 (6.4%) | 0 (0%) | 98 (3.9%) |  |  | 103 (3.9%) |
| Man: FSW-based channels | 22 (28%) | 10 (21%) | 588 (24%) |  |  | 620 (24%) |
| Woman: FSW-based channels | 14 (18%) | 16 (33%) | 655 (26%) |  |  | 685 (26%) |
| Man: Other delivery channels | 1 (1.3%) | 3 (6.3%) | 133 (5.3%) |  |  | 137 (5.2%) |
| Woman: Other delivery channels | 1 (1.3%) | 1 (2.1%) | 71 (2.9%) |  |  | 73 (2.8%) |
| **Age group** |  |  |  | 0.5 | 0.11 |  |
| 15-24 years or less | 27 (35%) | 21 (44%) | 1 116 (45%) |  |  | 1 164 (45%) |
| 25-34 years | 38 (49%) | 20 (42%) | 1 005 (40%) |  |  | 1 063 (41%) |
| 35 years or more | 13 (17%) | 7 (15%) | 368 (15%) |  |  | 388 (15%) |
| **Marital status** |  |  |  | 0.3 | 0.5 |  |
| single | 54 (69%) | 32 (67%) | 1 675 (67%) |  |  | 1 761 (67%) |
| divorced / separated / widowed | 6 (7.7%) | 2 (4.2%) | 89 (3.6%) |  |  | 97 (3.7%) |
| living with partner / married | 18 (23%) | 14 (29%) | 725 (29%) |  |  | 757 (29%) |
| **Educational level** |  |  |  | 0.079 | 0.09 |  |
| none / primary | 13 (17%) | 13 (27%) | 477 (19%) |  |  | 503 (19%) |
| secondary | 50 (64%) | 29 (60%) | 1 353 (54%) |  |  | 1 432 (55%) |
| higher | 15 (19%) | 6 (13%) | 659 (26%) |  |  | 680 (26%) |
| **First-time tester** |  |  |  | 0.2 | 0.228 |  |
| no | 40 (51%) | 25 (52%) | 1 472 (59%) |  |  | 1 537 (59%) |
| yes | 38 (49%) | 23 (48%) | 1 017 (41%) |  |  | 1 078 (41%) |

FSW: female sex workers, MSM: men having sex with men.

**Table S5.** Time between HIVST and confirmatory testing among phase 2 participants who did link to confirmatory testing, by reported number of lines and self-interpreted HIVST result

|  | Overall | 2 lines /reactive | 1 line /reactive | 2 lines /non-reactive | 2 lines /DK-R | DK-R / reactive |
| --- | --- | --- | --- | --- | --- | --- |
| less than a week | 18 (53%) | 12 (80%) | 0 (0%) | 0 (0%) | 6 (75%) | 0 (0%) |
| between 1 and 2 weeks | 4 (12%) | 1 (6.7%) | 0 (0%) | 2 (22%) | 1 (12%) | 0 (0%) |
| between 3 and 4 weeks | 2 (5.9%) | 1 (6.7%) | 0 (0%) | 0 (0%) | 1 (12%) | 0 (0%) |
| between 1 and 2 months | 7 (21%) | 1 (6.7%) | 0 (0%) | 5 (56%) | 0 (0%) | 1 (100%) |
| more than 3 months | 3 (8.8%) | 0 (0%) | 1 (100%) | 2 (22%) | 0 (0%) | 0 (0%) |
| Total | 34 (100%) | 15 (44.2%) | 1 (2.9%) | 9 (26.5%) | 8 (23.5%) | 1 (2.9%) |

DK: don't know. R: refuse to answer

**Table S6.**
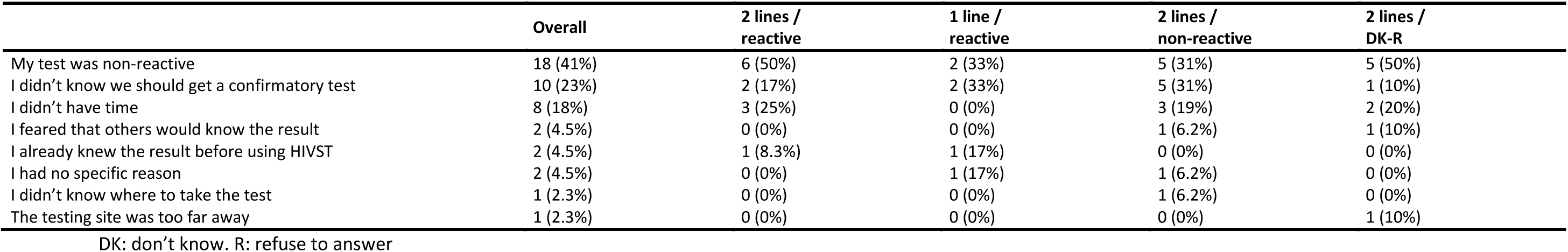
Main reason for not linking to confirmatory testing among phase 2 participants who did not link to confirmatory testing, by reported number of lines and self-interpreted HIVST result.

**Table S7.**
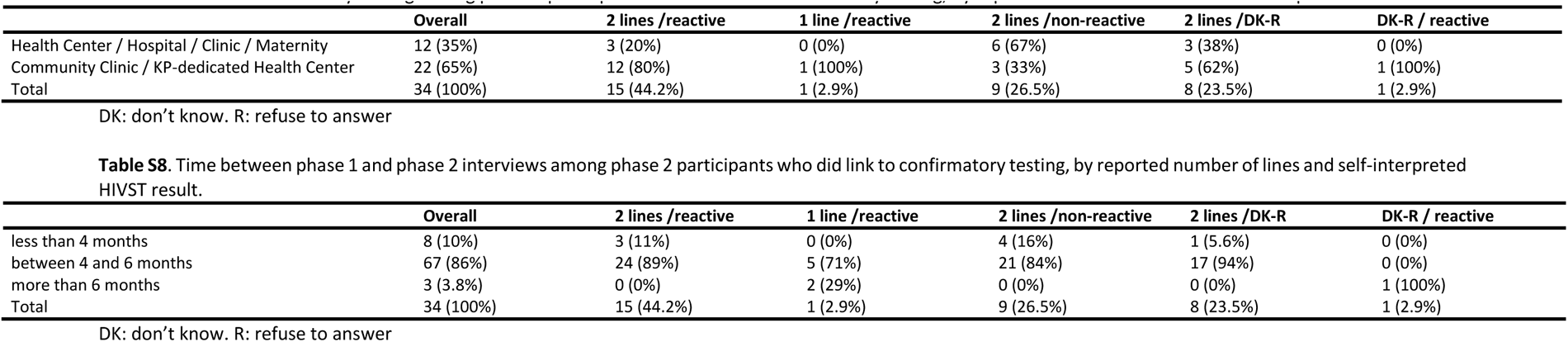
Place of confirmatory testing among phase 2 participants who did link to confirmatory testing, by reported number of lines and self-interpreted HIVST result.

**Table S8.**
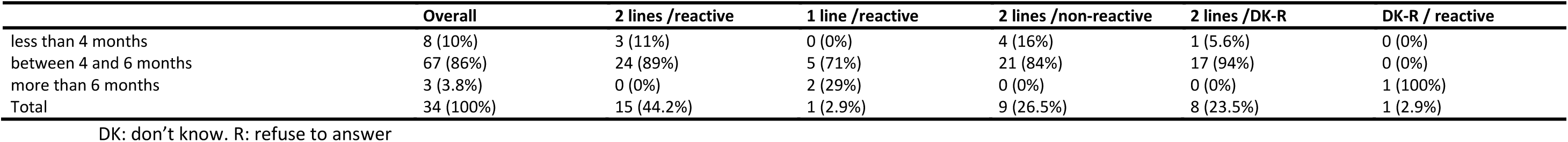
Time between phase 1 and phase 2 interviews among phase 2 participants who did link to confirmatory testing, by reported number of lines and self-interpreted HIVST result.

